# Long-term humoral response following simultaneous Delta and Omicron BA.1 co-infection

**DOI:** 10.1101/2022.10.28.22281636

**Authors:** Carla Saade, Bruno Pozzetto, Melyssa Yaugel Novoa, Bruno Lina, Stéphane Paul, Antonin Bal, Sophie Trouillet-Assant, Lyon COVID study group

## Abstract

To provide insight into the long-term immune response following bivalent vaccines, we sampled vaccinated patients simultaneously co-infected with Delta and BA.1. We reported that simultaneous exposure to the Delta and BA.1 S protein does not confer an additional immune advantage compared to exposure to the Omicron BA.1 S protein alone.

---

Bivalent vaccines containing two different sequences of the Spike (S) protein have been recently approved. First studies evaluating the short-term immune response after the administration of bivalent vaccines have reported contradictory results^1–3^. Moreover, the long-term immunity outcomes of this bivalent vaccine need further investigation, especially given the waning of the humoral response. Others and we have reported cases of simultaneous co-infection by the Delta and BA.1 variants, both have caused major COVID-19 epidemic waves worldwide^4,5^. The present report describes the long-term humoral response of patients co-infected with the variants Delta and BA.1 and aims at providing insights into that induced by bivalent vaccines.

We included two groups of fully vaccinated individuals with a breakthrough infection (BA.1 in the first group [n=9] and Delta and BA.1 co-infection in the second group [n=9]) as well as a third group of COVID-19-naïve individuals who received three doses of the BNT162b2 vaccine (n=9) (Table S1). Blood sampling was performed 6 months after breakthrough infection for infected individuals or after the third vaccine injection for COVID-19-naïve individuals (Table S1). Statistical analyses were performed using the GraphPad Prism® software (version 8; GraphPad software, La Jolla, CA, USA). Participants were included from two clinical studies registered on ClinicalTrial.gov (NCT05060939, NCT04341142). Written informed consent was obtained from all participants and approval was obtained from the regional review board in April 2020 (*Comité de Protection des Personnes Sud Méditerranée I*, Marseille, France; ID-RCB 2020-A00932-37; ID-RCB 2021-A01877-34).

There was no significant difference in anti-RBD IgG and anti-S1 IgA levels among individuals with a breakthrough infection caused by Delta and BA.1 or by BA.1 only; however these levels were 4.13 and 10.83 fold lower, respectively, among COVID-19-naïve individuals (Fig. 1A and 1B).

**Figure 1.**
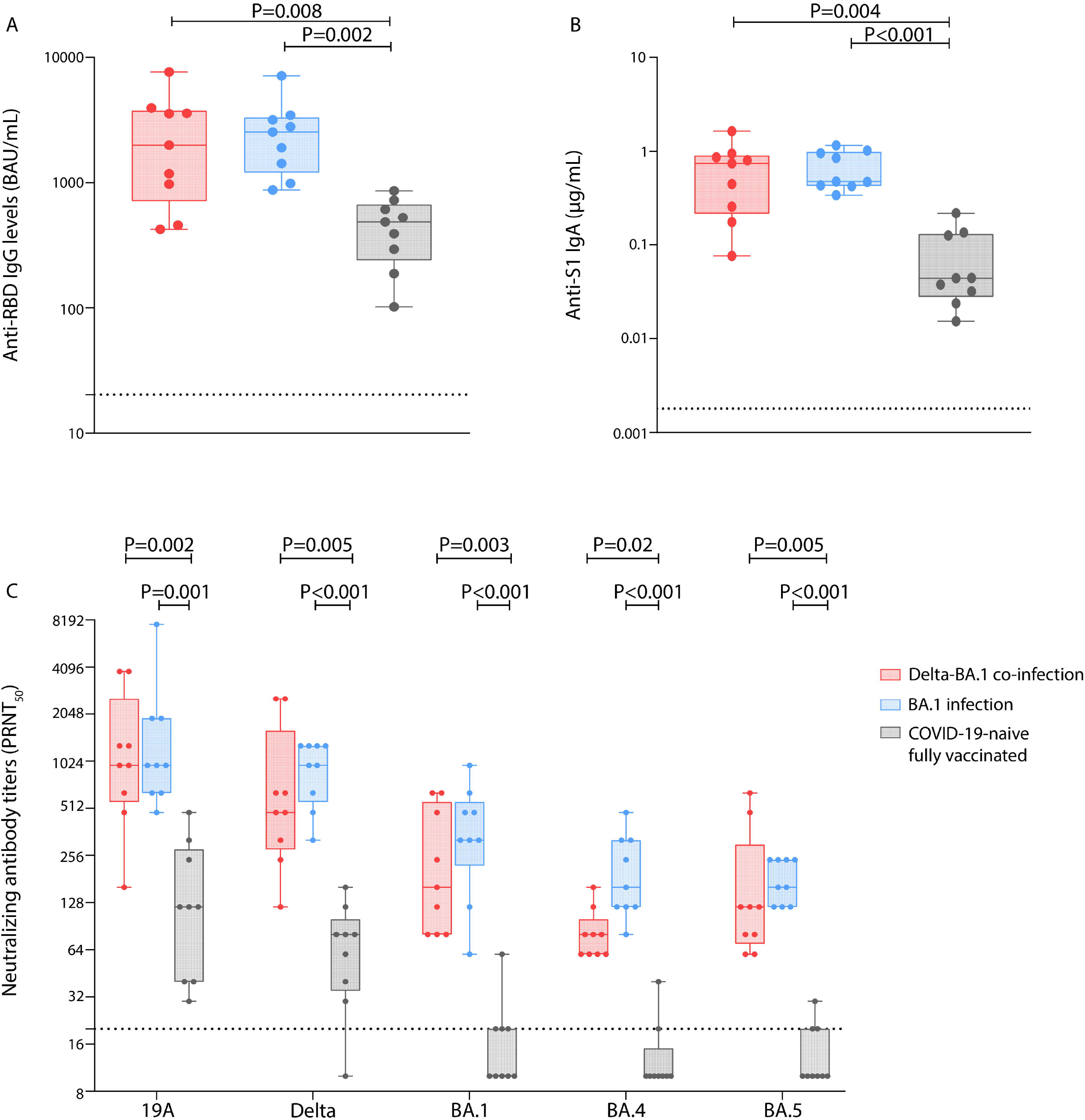
Humoral immune response 6 months after immunization in individuals vaccinated and then co-infected with Delta and BA.1, or vaccinated and then infected with BA.1, or in COVID-19-naïve fully-vaccinated individuals. Blood samples were obtained 6 months after infection or 6 months after the booster injection from 9 individuals in each group of patients. Anti-RBD IgG levels were measured using the bioMérieux Vidas SARS-CoV-2 IgG diagnosis kit according to manufacturer’s recommendations and expressed in binding antibody unit (BAU) /mL. The dotted line represents the threshold of positivity (≥ 20.33 BAU/mL) (**Panel A**). Anti-S1 IgA levels were measured using an ELISA test as previously described ^7^ and expressed in µg/mL. The dotted line represents the positivity threshold (≥ 0.0018 µg/mL) (**Panel B**). Neutralizing antibody titers against live SARS-CoV-2 isolates using a 50% plaque reduction neutralization test (PRNT_50_), as previously described ^7^. The isolates used for these experiments were 19A, Delta, Omicron BA.1, BA.4, and BA.5; their GISAID accession numbers are EPI_ISL_1707038, EPI_ISL_1904989, EPI_ISL_7608613, EPI_ISL_12396843, and EPI_ISL_12852091, respectively. The dotted line represents the positivity threshold (titer ≥ 20) (**Panel C**). Data are represented as box and whiskers plot; in each plot, the dots indicate individual samples, the upper and lower limits of the box plot represent the interquartile [IQR] range and the middle line represents the median. A Kruskal-Wallis test followed by Dunn’s multiple comparison tests were performed to assess differences between the three groups.

Neutralizing antibody titers were significantly lower among COVID-19-naïve individuals for all viral variants compared to BA.1-infected and Delta-BA.1-co-infected individuals. Compared to COVID-19-naïve individuals, the median neutralizing antibody titer of BA.1-infected individuals was 8-fold higher for the 19A isolate and 12-fold higher for the BA.5 isolate, and it was 8-fold higher for the 19A isolate and 16-fold higher for the BA.5 isolate for Delta-BA.1 co-infected individuals. Interestingly, no significant difference was observed between the two groups of individuals with a breakthrough infection (Fig. 1C).

Results reported herein showed an enhanced humoral response after breakthrough infections caused by one or two variants. These results confirmed the advantage conferred by so-called hybrid immunity, that has already been shown to induce a more potent long-term immune response against SARS-CoV-2 in comparison to that induced by vaccination only ^6^.

In addition, the absence of significant difference in anti-S total antibodies and neutralization capacity between individuals with a single or a dual breakthrough infection would suggest that a simultaneous exposure to the Delta and Omicron BA.1 variants does not confer an additional immune advantage in terms of humoral immunity. Our results suggest that a combined exposure to two different S proteins, such as in the case of bivalent vaccines, does not confer a long-term immune advantage in comparison to a sequential exposure to different S proteins. Studies conducted by Wang et al. and Collier et al. investigated the immune response following a bivalent vaccination in comparison to a monovalent vaccination ^2,3^. These studies showed no significant difference in neutralization capacity against Omicron subvariants between bivalent and monovalent vaccination. These results indicated the possibility of an immune imprinting in the context of anti-SARS-CoV-2 response, also known as original antigenic sin. This phenomenon is in reference to a limited immune response against a new antigen variant after an exposure to the initial antigen.

As our series was limited to cases of spontaneous infections with two different variants, studies comparing the long-term immune response induced after a challenge using a monovalent or a bivalent vaccine in COVID-19-naïve individuals are needed to evaluate the true benefit of the second strategy.

## Data Availability

All data produced in the present study are available upon reasonable request to the authors

## Acknowledgment

We thank all the staff members of the SARS-CoV-2 testing centre of the Hospices Civils de Lyon who contributed to the sample collection. We thank Cecile Barnel, the Clinical Research Associates, for her excellent work. We thank all the technicians from the virology laboratory whose work made it possible to obtain all these data. We thank all the members of the clinical research and innovation department for their reactivity (DRCI, Hospices Civils de Lyon). We thank Hélène Boyer (DRCI, Hospices Civils de Lyon) for her help in manuscript preparation. We thank all members of the Lyon Covid-ser study group: Jean-Baptiste Fassier, Nicolas Guibert, Dulce Alfaiate, Amélie Massardier-Pilonchery, Vanessa Escuret, Hélène Lozano, Laurence Josset, Bouchra Mokdad, Fanny Joubert and Camille Mena. Lastly, we thank all the participants for their involvement in this clinical study.

