## Supplemental Table 1 for "Long-term humoral response following simultaneous Delta and Omicron BA.1 co-infection"

**Supplementary Appendix**

### **Table S1:** Demographic and clinical characteristics of the participants

|  | Delta-BA.1 co-infection  (n=9) | BA.1  infection (n=9) | COVID-19-naïve individuals (n=9) |
| --- | --- | --- | --- |
| Female, n (%) | 3 (33) | 6 (66) | 6 (66) |
| Age, years, median [IQR] | 32 [23-40] | 49 [29-54] | 51 [48-64] |
| Delay between last immunization and sampling, days, median [IQR] | 203  [197-212] | 193  [208-166] | 179  [178-183] |
| Comorbidities, n (%) | 1 (11) | 1 (11) | 6 (66) |
| - hypertension, n (%) | 0 | 0 | 1 (11) |
| - diabetes, n (%) | 0 | 0 | 1 (11) |
| - hypothyroid, n (%) | 0 | 0 | 1 (11) |
| - vascular disease, n (%) | 0 | 1 (11) | 0 |
| - chronic lung disease, n (%) | 1 (11) | 0 | 2 (22) |
| - heterozygous sickle cell disease, n (%) | 0 | 0 | 1 (11) |
| Vaccination scheme |  |  |  |
| 3 injections of BNT162b2 | 3 (33) | 3 (33) | 9 (100) |
| 2 injections of BNT162b2 | 4 (44) | 4 (44) | 0 |
| Hybrid immunity | 2 (22) | 2 (22) | 0 |

IQR: interquartile range
